# Paired molecular characterization of repeat *Chlamydia trachomatis* infection among pregnant women in East London, South Africa

**DOI:** 10.64898/2026.11.20.26368559

**Authors:** Mildred Mandisa Mdingi, Brian Kullin, Hyunsul Jung, Ranjana M.S. Gigi, Chibuzor M. Babalola, Andrew Medina-Marino, Christopher M. Taylor, Christina A. Muzny, Nicola Low, Jeffrey D. Klausner, Remco P.H. Peters

**Affiliations:** Department of Medical Microbiology, University of Pretoria, Pretoria, South Africa; Research Unit, Foundation for Professional Development, East London, South Africa; Department of Pathology, University of Cape Town, Cape Town, South Africa; DST-NRF CAPRISA Centre of Excellence in HIV Prevention, Durban, South Africa; Institute of Social and Preventive Medicine, University of Bern, Bern, Switzerland; Keck School of Medicine, University of Southern California, Los Angeles, California, USA; Department of Epidemiology and Biostatistics, Michigan State University, Grand Rapids, Michigan, USA; Department of Medicine, Michigan State University, Grand Rapids, Michigan, USA; Department of Microbiology, Genetics, and Immunology, Louisiana State University, Health Sciences Center, New Orleans, Louisiana, USA; Division of Infectious Diseases, University of Alabama at Birmingham, Birmingham, Alabama, USA; Division of Medical Microbiology, Faculty of Medicine, University of Cape Town, Cape Town, South Africa

**Keywords:** *Chlamydia trachomatis*, *ompA* typing, repeat infection, pregnancy, South Africa

## Abstract

**Background:** Repeat *Chlamydia trachomatis* positivity after treatment may reflect persistent infection, reinfection, or residual nucleic acid. We compared paired *ompA* genotypes of *C. trachomatis* in pregnant women in South Africa and assessed macrolide resistance-associated mutations.

**Methods:** We analyzed paired baseline and follow-up specimens from 43 pregnant women with *C. trachomatis* detected at two study visits. We determined *ompA* genotypes by PCR amplification and Sanger sequencing and compared genotypes between visits. A fragment of the 23S rRNA gene was sequenced to identify selected macrolide resistance-associated mutations.

**Results:** At baseline, *ompA* typing was successful in 38/43 (88%) specimens and 36/43 (84%) at follow-up, with paired genotype results available for 32 women (74%). Genotypes E, J and D predominated at baseline and follow-up. The same genotype was detected at both visits in 22/32 women (69%), whereas 10/32 (31%) had a different genotype at follow-up. Genotype concordance was observed in 2/2 (100%) women with paired results at the test-of-cure visit, 13/16 (81%) assessed at 32 weeks gestation, and 7/14 (50%) assessed postnatally. No targeted macrolide resistance associated mutations were detected in specimens with successful 23S rRNA sequencing.

**Conclusions:** One-third of women with paired results had a different *ompA* genotype at follow-up, consistent with acquisition of a different strain after treatment. Concordant *ompA* genotypes may reflect persistence or reinfection with a strain sharing the same genotype, acquired from the same or a different partner. Macrolide resistance associated mutations were not detected. High-resolution typing methods would allow more detailed understanding of persistent *C. trachomatis* test positivity in this population.

## INTRODUCTION

*Chlamydia trachomatis* is a common bacterial sexually transmitted infection (STI) among pregnant women and is frequently asymptomatic. A systematic review reported a pooled *C. trachomatis* prevalence of 10.8% (95% CI:6.9-15.5) among pregnant women living in sub-Saharan Africa (1). This includes relatively high estimates from South Africa with recent studies reporting prevalence estimates of 20% to 24% among women attending antenatal care (ANC) in Western Cape, 11% in Kwazulu-Natal, 14.7% in the Eastern Cape, and 29.5% among pregnant women living with HIV in Tshwane (2-6). Untreated *C. trachomatis* infection during pregnancy may be associated with multiple adverse pregnancy outcomes including preterm birth, premature rupture of membranes, still birth and low birth weight (7). If present at the time of birth, *C. trachomatis* can cause neonatal conjunctivitis and pneumonia.

STI management among pregnant women in South Africa is complicated by repeat infections following treatment. In Tshwane, 27 of 102 (26.5%) pregnant women who returned for test of cure remained positive for *C. trachomatis* (8). Similarly, in Buffalo City, Eastern Cape Province, 8 of 65 (12%) women with baseline *C. trachomatis* infection who attended the test-of-cure visit had repeat-positive results. In the same study, repeat STI positivity was lower among women who reported that their partners had received treatment than among those whose partners had not received treatment (9). These findings highlight the challenge of preventing repeat *C. trachomatis* positivity during pregnancy.

A positive nucleic acid amplification test (NAAT) results following treatment may reflect different clinical scenarios (10). Treatment failure may occur due to poor adherence to treatment or severe gastrointestinal adverse events following oral treatment, or due to antimicrobial resistance (AMR), although this has not been document for *C. trachomatis* (11-13). Residual genetic material from *C. trachomatis* organisms may remain detectable in clinical samples after successful treatment, resulting in a positive NAAT result when testing is performed too soon after treatment (11, 14). Reinfection may occur through sexual contact with the same partner if that partner has not been treated, or through sexual contact with a new partner who has the infection (10). These scenarios require different clinical management to effectively cure the infection.

Molecular typing, including multi locus sequence typing (MLST) and whole genome sequencing (WGS) provides an opportunity to investigate some of the reasons for repeat *C. trachomatis* test positivity by comparing the genotype of strains identified at different time points. Conventional *ompA* typing has been widely used for epidemiological characterization of *C. trachomatis*, including investigation of repeat infection (15-18).

Despite the high prevalence of *C. trachomatis* infection among pregnant women in South Africa, the origins of repeat test positivity remain poorly understood. The objectives of this study were to compare *ompA* genotypes in paired baseline and follow-up *C. trachomatis*-positive specimens from pregnant women and to screen for selected macrolide resistance-associated mutations in the 23S rRNA gene.

## MATERIALS AND METHODS

### Description of parent studies and sample selection

The present study used stored specimens from two parent studies conducted between March 2021 and May 2024: a randomized, three-group trial evaluating STI testing approaches and adverse birth outcomes, and a prospective cohort study of genital infections, the vaginal microbiome and birth outcomes. Both studies recruited pregnant women from primary healthcare facilities in the Buffalo City Metropolitan Health District, Eastern Cape, South Africa. The parent study protocols have been published (19, 20).

In the parent studies, research staff collected multiple vaginal swabs for microbiological testing including a swab for Xpert® testing and a dry flocked swab (Copan, Brescia, Italy) that was stored at -20°C in the Department of Microbiology at the University of Pretoria for further molecular analysis. Baseline specimens were obtained from all women and tested on-site for *C. trachomatis* using the Xpert® CT/NG assay (Cepheid, Sunnyvale, CA, USA). Women with a positive test result for *C. trachomatis* received directly observed treatment with a single 1g oral dose of azithromycin. Repeat sample collection was determined by the parent study: test-of-cure (TOC) at 21 days post treatment (randomized controlled trial), 32 weeks of gestation (randomized controlled trial, cohort study), or at the 3-6 days first postnatal visit (randomized controlled trial).

### Current study design

This analysis included women who tested positive for *C. trachomatis* at their baseline visit and who had a positive test result at a follow-up study visit as described above. Stored paired vaginal swab specimens collected at baseline and follow-up were retrieved and processed for molecular typing.

The study was approved by the University of Pretoria Faculty of Health Sciences Research Ethics Committee (UP HREC reference 42/2024). All laboratory procedures were conducted at the Department of Medical Microbiology, University of Pretoria.

### DNA extraction

Total genomic DNA was extracted from stored specimens using the High Pure PCR Template Preparation Kit (Roche Diagnostics GmbH, Germany) according to the manufacturer’s instructions. The swab samples were prepared for extraction by vortexing in 1 mL of sterile Tris-EDTA buffer (compromising Tris(hydroxymethyl) aminomethane and ethylenediaminetetraacetic acid; Sigma-Aldrich, USA) for 5 minutes, after which they were centrifuged (5417R, Eppendorf, Germany) at 13 000 x g for 5 minutes and the resulting pellet resuspended in 200 μL of sterile phosphate-buffered saline buffer (Sigma-Aldrich, USA). The resuspension was subsequently processed according to manufacturer’s instructions. Extracted DNA samples were stored at −20°C until further analysis.

### *ompA* genotyping

The *ompA* gene was amplified by nested PCR using previously published primer sequences (21). PCR was performed on a SimpliAmp^TM^ Thermal Cycler (ThermoFisher Scientific Inc., USA). The PCR amplification of the outer target region was conducted in a final volume of 25 μL containing CelTaq HS DNA Polymerase and the corresponding buffer (Celtic Molecular Diagnostics, South Africa), outer primers at a final concentration of 0.8 μM each, and 2 μL of extracted DNA. The amplification protocol consisted of an initial denaturation at 95°C for 3 minutes, followed by 35 cycles of denaturation at 93°C for 30 seconds, annealing at 59°C for 30 seconds, and extension at 72°C for 1 minute. A final extension step was performed at 72°C for 5 minutes. For the PCR of the inner target region, 2 μL of the outer PCR product was used as the template, and amplification was carried out for 30 cycles using the same cycling conditions. PCR products were assessed by electrophoresis on a 1% (w/v) agarose gel. Amplicons were then purified and sequenced in both directions by Sanger sequencing at Inqaba Biotechnical Industries, South Africa.

### Detection of 23S rRNA macrolide resistance-associated mutations

The 23S rRNA gene region encompassing positions A2059 and C2611, which are associated with macrolide resistance in *Mycoplasma genitalium*, *Neisseria gonorrhoeae* and *Treponema pallidum* but have not been reported in *C. trachomatis*, was amplified by PCR from extracted DNA using previously published primer sequences and cycling conditions (13). Each PCR reaction contained CelTaq HS DNA Polymerase and its corresponding buffer (Celtic Molecular Diagnostics, South Africa), the Ct23S-M1F and Ct23S-M2-R primers at 0.4 μM each, and 2 μL of template DNA. The expected amplicon size was 1,090 bp. PCR products were assessed by electrophoresis on a 1% (w/v) agarose gel. Amplicons were subsequently purified and subjected to four Sanger sequencing reactions per amplicon at Inqaba Biotechnical Industries (Pty) Ltd, South Africa, using two forward primers (Ct23S-M1-F and Ct23S-M2-F) and two reverse primers (Ct23S-M1-R and Ct23S-M2-R) to obtain bidirectional sequence reads. Sequence data were analysed for macrolide resistance associated mutations in the targeted 23S rRNA region.

### Bioinformatics analysis

The *ompA* genotype was determined using a curated offline reference database generated from *ompA* gene sequences extracted from previously characterised whole genome sequences representing known *ompA* genotypes (22). Raw sequence data were edited using BioEdit,(23) and multiple sequence alignments were generated with ClustalW implemented in MEGA v12.0.10 prior to genovar assignment (24). The final *ompA* alignment was also used to construct a maximum likelihood phylogeny using IQ-TREE2 v2.4.0,(25) using a best fit model selected by the ModelFinder option of the program with 1000 bootstrap replicates (26).

For the analysis of macrolide resistance associated mutations positions A2059 and C2611 within the 23S rRNA gene, sequence chromatograms were analysed using Staden Package version 2.0. Edited sequences were compared with reference sequences using BLAST (27, 28). Sequence quality was assessed using Phred scores, and only sequence reads with an average Phred score >20 were included in the analysis.

### Statistical analysis

Categorical variables were summarised using frequencies and percentages, while continuous variables were summarised using medians and ranges. Genotype concordance was calculated as the proportion of women with the same *ompA* genotype at baseline and follow-up among those with successful paired genotype results.

## RESULTS

This analysis included samples from 43 women with a positive *C. trachomatis* test result at their baseline visit and at a follow-up visit. Of these, 32 (74%) participated in the randomized trial and 11 (26%) in the prospective cohort study. Repeat *C. trachomatis*-positive specimens were collected at the test-of-cure visit 21 days after treatment in 8 women, at the 32 weeks visit in 20 women, and at the postnatal visit in 15 women. At baseline, the median age of participants was 23 years (range 18-40 years), 7/43 (16%) had a history of recent STI treatment, and 12/43 (28%) were living with HIV (Table 1).

**Table 1.** Baseline characteristics of study population (n=43)

|  | **Number (%) or median (range)** |
|---|---|
| Median age in years | 23 (18-40) |
| Employment status |  |
| Employed (formal or self) | 12 (28%) |
| Unemployed | 31 (72%) |
| History of STI treatment |  |
| Yes | 7 (16%) |
| No | 36 (84%) |
| HIV status |  |
| HIV-positive | 12 (28%) |
| HIV-negative | 30 (70%) |
| Unknown | 1 (2.3%) |
| Relationship status |  |
| Married | 3 (7.0%) |
| Steady partner | 38 (88%) |
| Steady and casual partner(s) | 1 (2.3%) |
| Casual partner(s) | 1 (2.3%) |
| Condom use during last sex act |  |
| Yes | 5 (12%) |
| No | 38 (88%) |

**Note.** STI, sexually transmitted infection; HIV, human immunodeficiency virus.

The *ompA* typing was successful in 38/43 (88%) of baseline specimens and 36/43 (84%) of follow-up specimens. Genotypes E (n=10), J (n=10), and D (n=9) were most frequently identified at baseline (Supplementary Table 1). At follow-up, the frequency of serovars was D (n=12), J (n=9), and E (n=7) (Supplementary Table 1). There was no apparent clustering of samples by site or participant characteristics, including HIV status, history of STI treatment, or relationship status (Figure 1).

**Figure 1.**
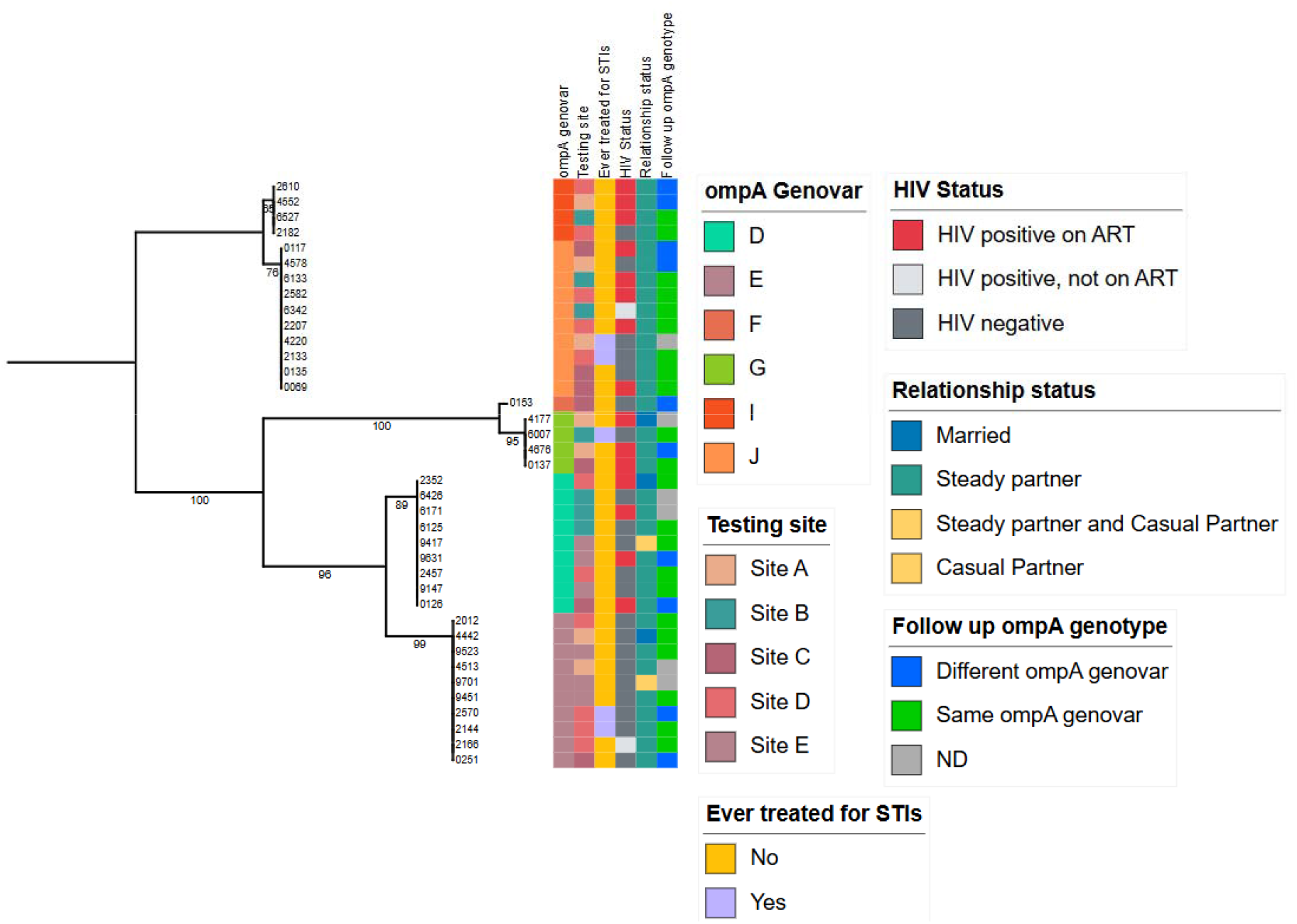
Maximum-likelihood phylogeny of *Chlamydia trachomatis ompA* sequences at baseline annotated by *ompA* genovar, participant characteristics, and follow-up *ompA* genotype concordance. Bootstrap support values are shown at internal nodes.

Paired genotype data for baseline and follow-up visit were available for 32/43 (74%) women. Genotyping results varied by follow-up visit: paired genotypes were obtained for 2/8 (25%) women at the test-of-cure visit at 21 days after treatment, 16/20 (80%) at 32 weeks’ gestation and 14/15 (93%) at the postnatal visit. The same *ompA* genotype was identified in baseline and follow-up samples from 22/32 (69%) women while 10/32 (31%) had a different *ompA* genotype at the follow-up visit (Table 2). Looking at timing of follow-up, the same genotype was identified in 2/2 (100%) women at the test-of-cure visit at 21 days after treatment, 13/16 (81%) among women at 32 weeks’ gestation, and 7/14 (50%) among women at their postnatal visit (Figure 2).

**Figure 2.**
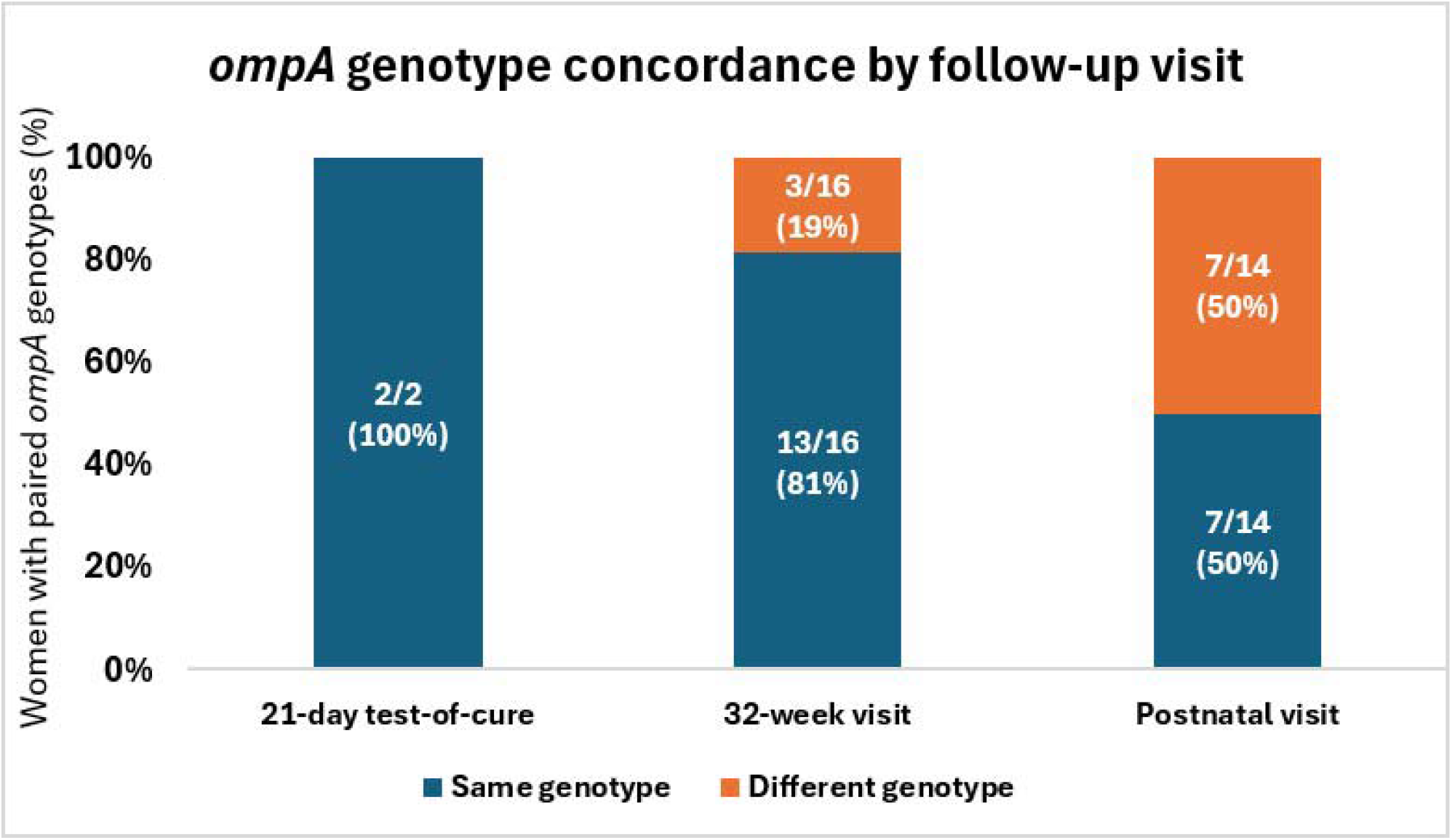
*ompA* genotype concordance between paired samples collected at follow-up visits (n=32)

**Table 2.** Characteristics of women with different *ompA* genotype *Chlamydia trachomatis* result between baseline and follow-up visit (n=10)

| **Study site** | **Patient** | ***ompA* baseline** | ***ompA* follow-up** | **Employment status** | **Relationship status** | **HIV status** | **Condom use last sex** |
|---|---|---|---|---|---|---|---|
| A | 1 | I | G | Unemployed | Steady partner | Positive, on ART | Yes |
| A | 2 | J | F | Unemployed | Steady partner | Negative | No |
| A | 3 | G | D | Employed | Steady partner | Positive, on ART | No |
| C | 4 | J | G | Unemployed | Steady partner | Negative | No |
| C | 5 | D | G | Unemployed | Steady partner | Negative | No |
| C | 6 | F | D | Unemployed | Steady partner | Positive, on ART | No |
| C | 7 | E | D | Unemployed | Steady partner | Negative | No |
| D | 8 | E | D | Unemployed | Steady partner | Negative | No |
| D | 9 | I | D | Unemployed | Steady partner | Positive, on ART | No |
| E | 10 | D | J | Unemployed | Steady partner | Positive, on ART | No |

**Note.** HIV, human immunodeficiency virus; ART, antiretroviral therapy.

In exploratory comparisons, women with a different genotype at follow-up were more likely to be unemployed (9/10), than employed ( (14/22, odds ratio, OR 5.1; 95% CI 0.55-48.4, p=0.15) and living with HIV (5/10) than not (6/22, OR 2.7; 95% confidence interval, CI 0.56-12.6, p=0.21) than to women with the same genotype at follow-up. There was no difference in partner types between the groups (Supplementary Table 2). Interpretable 23S rRNA sequences were obtained from 36/43 (83.7%) baseline specimens and 32/43 (74.4%) follow-up specimens. No macrolide resistance associated mutations were detected at positions A2059 or C2611 in the successfully sequenced specimens.

## DISCUSSION

This study used *ompA* genotyping of paired samples to investigate repeat *C. trachomatis*-positive results among pregnant women in South Africa. Approximately two-thirds of women with successful genotyping had concordant genotypes at both visits. Detection of a different genotype is consistent with acquisition of a new infection. However, this finding does not necessarily confirm reinfection, as mixed *C. trachomatis* genovar infections have been documented. Using high-throughput sequencing, Zhao, et al. (29) identified mixed-genotype infections in 20 of 303 (6.6%) typed cervical specimens, demonstrating that more than one genovar can be present in a single specimen. Therefore, a genovar detected at follow-up but not identified at baseline may have been present in the initial specimen but not detected by the genotyping approach used. In contrast, a concordant genotype does not distinguish persistence of the baseline infection from reinfection with the same strain or another strain sharing the same *ompA* genotype. No macrolide resistance associated mutations were detected in relation to persistent test positivity.

In our study, concordant ompA genotypes were observed in 22/32 (69%) paired specimens. This proportion was numerically higher than the 20% concordance reported among the 35 paired post-treatment specimens from the STD clinic clients in the United States and the 59% (10/17) reported among young heterosexual people in the Netherlands (16, 18). Differences in study populations, pregnancy status, circulating genotypes, follow-up intervals, treatment and partner-management approaches may contribute to these differences (16, 18). The genotype distribution in our study was broadly consistent with previous South African findings, with genovars D and E commonly identified. Genovar J was also frequent in our population. It has also been reported in other South African populations with variation across geographical settings and study population. Versteeg, et al. (17) described urogenital *C. trachomatis* strains among women in Limpopo Province, while Mofolorunsho, et al. (30) reported genotypes among men who have sex with men in KwaZulu-Natal. More recently Dabee, et al. (15) reported differences in genovar distributions between adolescent girls and young women in the Western Cape and Gauteng provinces.,

Genotype concordance varied across follow-up visits, from 100% at the 21 days post-treatment visit, to 81% among women assessed at 32 weeks’ gestation and 50% among women assessed postnatally. A longer interval post-treatment provides more opportunity for sexual re-exposure and could therefore increase the probability of detecting a new infection. However, these comparisons involved small non-random subgroups and only 2 women had paired results at the 21-day test-of-cure visit.

All 10 women with genotype discordance at follow-up reported a steady partner and nine reported no condom use during their most recent sexual encounter. These findings indicate the potential for sexual re-exposure during the interval between baseline and follow-up but cannot establish the source of infection. cannot establish when transmission occurred or identify its source. Partner treatment may provide additional context because exposure to an untreated partner is an important contributor to repeat infection. In a related analysis of pregnant women with repeat STI positivity, including *C. trachomatis*, *N. gonorrhoeae* and *T. vaginalis*, 48/108 (44%) reported that their partners had received treatment. Repeat STI positivity was less frequent among women reporting partner treatment than among those reporting untreated partners (4% vs 27%, respectively (9).

Sequencing of the 23S rRNA gene did not identify macrolide resistance associated mutations at positions A2059 or C2611 in any of the 43 women. These findings do not establish azithromycin susceptibility but do suggest that these specific macrolide resistance associated mutations did not contribute to persistent test positivity.

This study has several strengths. It used paired specimens collected before and after azithromycin treatment from pregnant women in a setting with a high burden of *C. trachomatis* infection allowing for within-participant comparison of genotypes. In addition, data on follow-up timing and selected partner and behavioural characteristics was included. Important study limitations include the small sample size and availability of paired *ompA* results for only 32/43 women. Follow-up occurred at different time intervals as determined by the parent studies. Mixed infections may not have been identified by this sequencing method, and *ompA* did not provide strain-level resolution, for which MLST or WGS would have been required. We also did not assess bacterial viability and therefore could not distinguish active infection from residual DNA detection

In conclusion, paired *ompA* typing showed that repeat *C. trachomatis* positivity after treatment included both genotype-concordant and genotype-discordant infections. Detection of a different genotype at follow-up suggests the presence of a new infection, highlighting the importance of prevention interventions. On the other hand, concordant genotypes could not distinguish persistent infection from reinfection but do support comprehensive partner management strategies. Use of higher-resolution molecular methods, such as MLST, WGS and targeted deep sequencing of additional variable genomic regions, could improve strain discrimination and detection of mixed-genovar infections beyond *ompA* genotyping alone. These approaches, combined with standardized follow-up and detailed treatment, partner-treatment sexual exposure information, are needed to better distinguish reinfection from persistent infection and further investigate the reasons for repeat *C. trachomatis* positivity during pregnancy.

## Data Availability

All data supporting the findings of this study are presented in the manuscript and its supplementary materials.

## Summary

Paired *ompA* genotyping identified concordant and discordant genotypes among women with repeat *Chlamydia trachomatis* positivity, but higher-resolution typing is needed to distinguish persistent infection from reinfection.

## Ethics

The study received ethical approval from the Research Ethics Committee of the Faculty of Health Sciences, University of Pretoria (reference number: 42/2024). The study was conducted in accordance with applicable ethical guidelines and regulations.

## Acknowledgments

We would like to acknowledge the support provided by the Foundation for Professional Development Research Unit and the Department of Medical Microbiology, University of Pretoria.

## Contributors

MMM conceptualised the study, conducted the laboratory work, performed bioinformatics and data analysis, and drafted the manuscript. HJ provided laboratory supervision and technical support for the molecular analyses. BK contributed to the bioinformatics and sequence analyses and critically reviewed and revised the manuscript. RMSG, CMB, AMM, CMT, CAM, NL and JDK contributed to the interpretation of findings and critically reviewed and revised the manuscript. RPHP provided overall supervision, contributed to the study design and interpretation of findings, and critically reviewed and revised the manuscript. All authors reviewed and approved the final manuscript.

