## Supplementary Table 1 for "Paired molecular characterization of repeat *Chlamydia trachomatis* infection among pregnant women in East London, South Africa"

**Supplementary Table 1. Frequency of *ompA* genotypes of *Chlamydia trachomatis* detected in vaginal swab samples from pregnant women in East London, South Africa (n=43)**

| **Genotype** | **Number (%) at baseline visit** | **Number (%) at follow-up visit** |
| --- | --- | --- |
| A | - | - |
| B | - | - |
| C | - | - |
| D | 9 (21%) | 12 (28%) |
| E | 10 (23%) | 7 (16%) |
| F | 1 (2.3%) | 1 (2.3%) |
| G | 4 (9.3%) | 5 (12%) |
| H | - | - |
| I | 4 (9.3%) | 2 (4.7%) |
| J | 10 (23%) | 9 (21%) |
| K | - | - |
| L | - | - |
| Typing unsuccessful | 5 (12%) | 7 (16%) |
