## Supplementary Table 2 for "Paired molecular characterization of repeat *Chlamydia trachomatis* infection among pregnant women in East London, South Africa"

**Supplementary Table 2. Exploratory comparison of participant characteristics with the same versus a different *ompA* genotype at baseline and follow-up visit (n=32)**

| **Characteristic** | | **Same genotype (n=22)** | **Different genotypes (n=10)** |
| --- | --- | --- | --- |
| Facility | |  |  |
|  | A | 1 (4.5%) | 3 (30%) |
|  | B | 5 (22.7%) | 0 |
|  | C | 3 (13.6%) | 4 (40%) |
|  | D | 9 (40.9%) | 2 (20%) |
|  | E | 4 (18.1%) | 1 (10%) |
| Employment status | |  |  |
|  | Employed (formal or self) | 8 (36.3%) | 1 (10%) |
|  | Unemployed | 14 (63.6%) | 9 (90%) |
| Relationship status | |  |  |
|  | Married | 2 (9.1%) | 0 |
|  | Steady partner | 19 (86.4%) | 10 (100%) |
|  | Casual partner(s) | 1 (4.5%) | 0 |
| HIV status | |  |  |
|  | Positive | 6 (27.2%) | 5 (50%) |
|  | Negative | 16 (72.7%) | 5 (50%) |
| Condom use last sex act | |  |  |
|  | Yes | 3 (13.6%) | 1 (10%) |
|  | No | 19 (86.3%) | 9 (90%) |
